# Added Value and Clinical Significance of Nonlinear Variability Indices of Walking Stride Interval in Neurodegenerative Diseases

**DOI:** 10.1101/2020.01.31.20019455

**Authors:** Dierick Frédéric, Vandevoorde Charlotte, Chantraine Frédéric, White Olivier, Buisseret Fabien

## Abstract

Though self-paced walking is highly stereotyped, the stride interval fluctuates from one stride to the next around an average value with a measurable statistical variability. In clinical gait analysis, this variability is usually assessed with indices such the standard deviation or the coefficient of variation (CV). The aim of this study is to understand the added value that nonlinear indices of walking stride interval variability, such as Hurst exponent (H) and Minkowski fractal dimension (D), can provide in a clinical context and to suggest a clinical significance of these indices in the most common neurodegenerative diseases: Parkinson, Huntington, and amyotrophic lateral sclerosis. Although evidence have been accumulated that the stride interval organization at long range displays a more random, less autocorrelated, gait pattern in neurodegenerative diseases compared with young healthy individuals, it is today necessary to recompute CV, H, and D indices in a unified way and to take into account aging impact on these indices. In fact, computed nonlinear indices of variability are mainly dependent on stride interval time series length and algorithms used, making quantitative comparisons between different studies difficult or even impossible. Here, we recompute these indices from available stride interval time series, either coming from our lab or from online databases, or made available to us by the authors of previously conducted research. We confirm that both linear and nonlinear variability indices are relevant indicators of aging process and neurodegenerative diseases. CV is sensitive to aging process and pathology but does not allow to discriminate between specific neurodegenerative diseases. D shows no significative change in pathological groups. However, since H index is correlated with Hoehn & Yahr scores and significantly lowered in patients suffering from Huntington’s disease, we recommend it as a relevant supplement to CV.

## Introduction

Since the seminal work of Graham Brown [1] on how rhythmic locomotor behaviors are controlled in mammals, research along this line has continued to expand and is now a priority topic in disciplines as diverse as physiology, neurosciences and biomechanics. Natural – even automatic – processes are always accompanied with some randomness. More recently, the investigation of the variability of those behaviors has revealed, on the one hand, significant highlights suggestive of underlying mechanisms and control and, on the other hand, general lessons on motor variability: “More variable does not mean more random, and more controllable does not mean more deterministic” [2].

For more than 20 years, researchers have been interested in the long-term structure of walking, and more particularly in the variability of stride interval (SI). Though human walking is highly stereotyped, the SI fluctuates from one stride to the next around an average value with a measurable statistical variability. It appears that the long-term SI pattern, during a walk for about ten minutes, is auto-correlated: the temporal characteristics of a step are strongly dependent on the temporal characteristics of the previous steps [3, 4]. During the 1990s, Hausdorff and his collaborators [3, 4] proposed a framework for analyzing temporal dynamics of walking SI over a consistent number of strides (above 500) which has since been successfully used in different physiological and pathological contexts such as performing a dual task during walking [5, 6], studying aging process during walking [4, 7, 8], investigating the influence of Parkinson’s disease [9–11] or peripheral neuropathy on walking [12].

Information about the variability of SI are easily provided by linear tools. Nonlinear goes one step beyond by assessing its complexity and predictability [13]. It is challenging to diagnosing neurodegeneration at an early stage. Yet, it is also critical since the sooner an impairment is identified, the larger the likelihood of efficient intervention. We therefore believe that the assessment of these two complementary indicators of movement variability could be of major practical importance for the clinician.

The mathematical tools preferably used to quantify structures based on their variability belong to nonlinear analysis [14], of which fractal analysis is a part. The factor that has been mostly used in the long-term structure discussed above is the Hurst’s exponent (H), originally developed in the field of hydrology [15]. This index assesses the predictability of SI time series. Put differently, it assesses whether its dynamics is autocorrelated or random. The Minkowski’s fractal dimension (D) of SI time series has also been proved to be a reliable estimator of complexity over time, see *e*.*g*. [13]. Where does the autocorrelations find its origins in the walking cycle ? What are the hidden physiological mechanisms that drive behaviors ? Clues or answers to these fundamental questions could be provided by genuine analyses of indexes such as H or D. However, the calculation methods used in the aforementioned studies lack homogeneity mainly in terms of the different algorithms used, which can lead to biases in their interpretation, and making quantitative comparisons between different studies hazardous.

There is therefore a necessity to process and analyze all available data recorded in different studies by adopting a unified protocol. Our aim is to assess the influence of aging in healthy subjects and of common neurodegenerative diseases – Parkinson’s and Huntington’s diseases and amyotrophic lateral sclerosis – on linear and nonlinear walking SI variability indexes, using either time series recorded in our lab, available in online databases or made available to us by the authors of previous research. The two main questions that will be addressed in this research work are the following. First, are walking SI variability indices able to differentiate healthy subjects from patients, or even better provide information about the neurodegenerative disease involved and its progress? The current literature already suggests an affirmative answer to this question in the case of Parkinson’s disease [9, 10], but these indexes still need to be explored in other degenerative diseases. Second, is a clinical use of the walking SI variability indices possible, specifically in terms of complexity and predictability of movement variability ? It has already been suggested that “healthy” biological systems are characterised by an optimal variability, observable through physiological signals, be it electrocardiogram [8] or more generic time series coming from motion analysis [16]. Along the lines of that framework we focus on the peculiar case of SI time series, with the hypothesis that any departure from an optimal variability defined by the behavior of healthy individuals may reveal a possible dysfunction to be identified.

## Materials and Methods

### Time series and variability assessment

SI time series are measured in individuals walking at self-selected pace during a typical 8-minute interval. Time series come from existing databases, personal data and data from authors of other previously published studies, all of which met standards of informed consent and ethical oversight. Table 1 presents a summary of the data we have gathered. We refer the interested reader to the cited references for more details about the experiments in which these time series were first obtained. Figure 1 depicts typical SI over time. One can readily observe that each time series fluctuates around an average value, and that the dynamics of these fluctuations is nontrivial. This dynamics is assessed by resorting to the methodology developed in [13], that we summarize below.

**Table 1.**
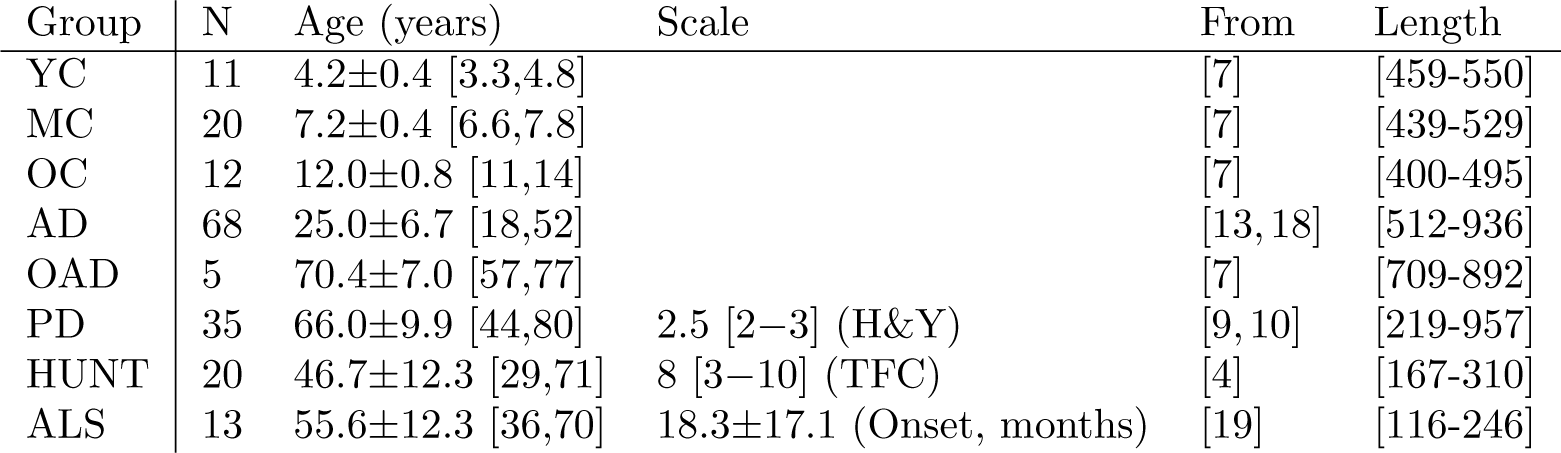
Summary of the different groups analysed in the present work. Data come from our own lab, from the free online PhysioNet Databases [17], or were given by the first author of [10]. For each group we display the number of time series available, the age of participants/ patients under the form mean±SD, and give the minimal and maximal ages between brackets. The minimal and maximal time series length in each group are given between brackets in the last column. We also summarize results from disability scales under the form median [Q1–Q3] when applicable. Healthy groups: Young Children (YC), Middle Children (MC), Old Children (OC), Adults (AD), Old Adults (OAD). Pathological groups: Parkinson’s disease (PD), Huntington’s disease (HUNT), amyotrophic lateral sclerosis (ALS). Disability scales: Modified Hoehn & Yahr (H&Y) for PD, Total Functional Capacity (TFC) for HUNT, number of months since first diagnosis (Onset) for ALS.

**Fig 1.**
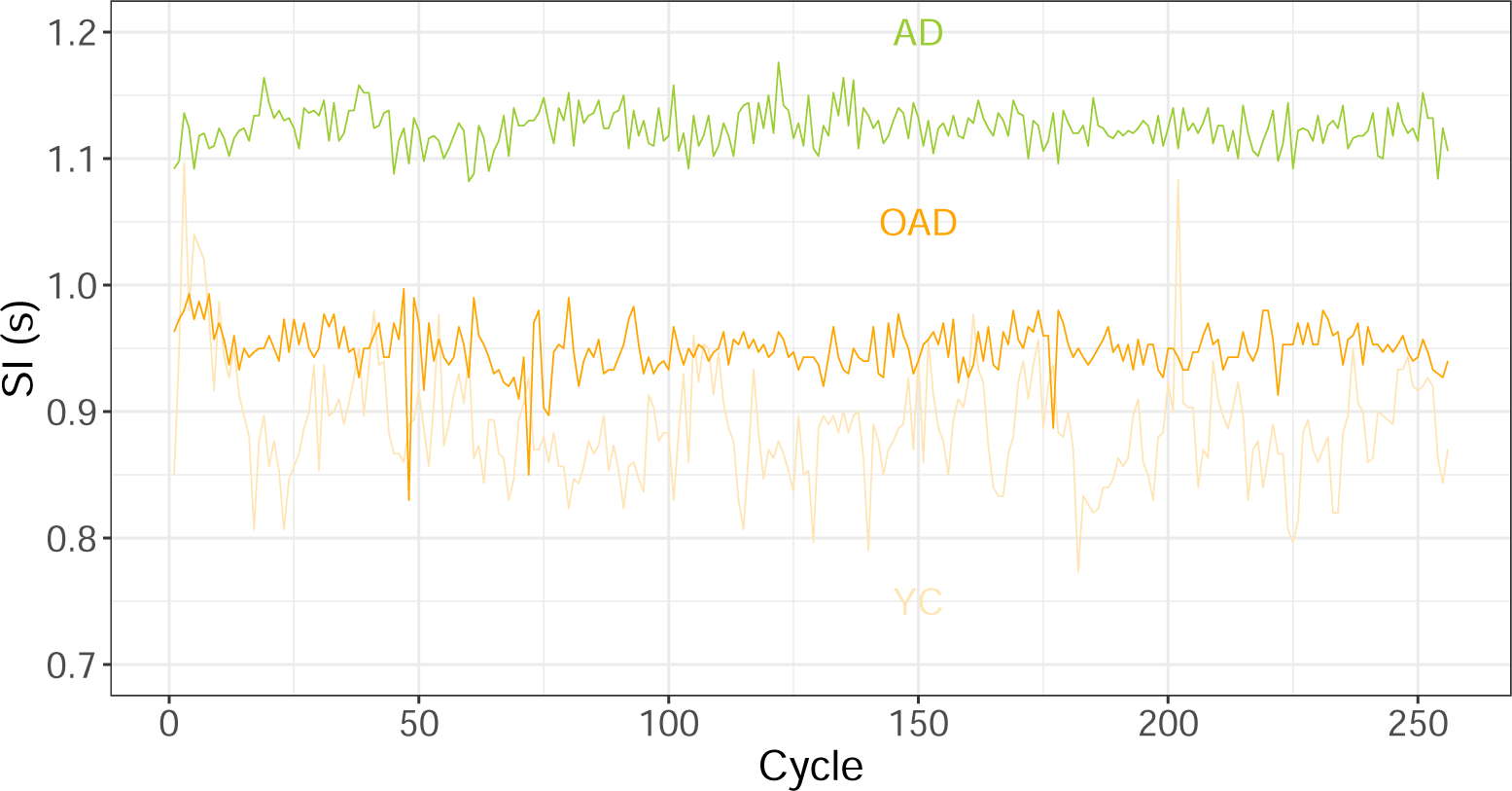
Typical SI time series for three healthy subjects of different ages: a young child (YC), an adult (AD) and an old adult (OAD).

Our data set consists in SI time series, denoted as ***T*** and containing the durations *T*_*i*_ of the successive cycles. The time series gathered are of variable length, in particular because of the small walking speed of patients suffering from neurodegenerative diseases. As shown in [20], the computed value of variability indices may significantly differ from their exact, asymptotic, value when time series are shorter than 256 points. We therefore choose to truncate time series longer than 256 SI to their last 256 points and to keep unchanged the shorter time series in order to reduce bias associated to time series lengths.

We first computed the average value *E*(***T***) and named it SI in the rest of this work. Then we computed three variability indicators: (1) the coefficient of variation, CV= *SD*(***T***)*/*SI, which yields the relative amplitude of the SI fluctuations around the mean value. It is worth noticing that CV does not provide any information on the temporal structure itself of these variations. Instead, these are quantified by the two independent indices D and H. (2) The Minkowski fractal dimension, D, is computed thanks to the box-counting method: if *N* (*ϵ*) is the number of square boxes of size *ϵ* needed to cover the plot of ***T***, then *N* (*ϵ* → 0) *ϵ*^−D^. D ranges from 1 to 2: The closer D to 2, the more important the SI relative fluctuations from one cycle to another. Intuitively, D measures the “apparent roughness” of ***T***, that we also call complexity. (3) The Hurst exponent H is computed by using the Detrended Fluctuation Analysis with a linear detrending (higher-order polynomial detrending is sometimes used in other similar studies). Computational details may be found in [21], but the main steps are as follows. The cumulated time series ***Z*** computed from ***T*** are divided into windows of length *l*. A local least squares linear fit is performed for each window and the fluctuation function *F* (*l*) is computed. The asymptotic relation *F* (*l* → ∞) ∝ *l*^H^ leads to H. A random process is close to H = 0.5, while long-range autocorrelated processes are such that 0.5 < H ≤ 1 (H > 1 for unstable processes) in which an increase in SI is likely to be followed by another increase in SI at long range, and similarly for a decrease. Time series with 0 < H < 0.5 are anticorrelated; this case is typically not encountered in SI time series. H is regarded as a predictability index. Strongly autocorrelated dynamics are such that the time series value at a given time is strongly dependent of its previous values, hence predictable.

Computation of SI, CV, H and D has been performed with R software (v.3.4.2) – packages *dfa* and *fractaldim*.

### Data analysis

We have first compared AD, PD, HUNT and SLA groups through a one-way ANOVA with significance level set at *p* = 0.05. In case of significant group effect, a Holm-Sidak *post-hoc* was used for comparison of the pathological groups (PD, HUNT, SLA) to the AD group seen as a control condition. Kruskal-Wallis test with Dunn’s method was used if normality test failed. ANOVAs were performed with SigmaPlot software (v.11.0,Systat Software, San Jose, CA).

Then we have computed Spearman’s correlation coefficients, *ρ*, between the different parameters and subject’s ages for all groups, all healthy subjects (YC, MC, OC, AD, OAD) having been gathered into the Healthy group. Within the Healthy group, *ρ* has been computed between the parameters themselves. In the pathological groups (PD, HUNT, ALS), *ρ* has been computed between the different parameters and disability scores. We consider a correlation as significant if "*ρ*| > 0.2 and *p <* 0.05 following Guilford lines [22]. Correlation between parameters have been further assessed by performing separated Principal Component Analysis (PCA) for Healthy and pathological groups.

Finally, we have compared the Healthy and pathological groups via a nonparametric ANCOVA [23] in order to “remove” the (nonlinear) age-dependence of parameters from the comparison. Significance level was set to *p* = 0.05. Spearman’s correlation coefficients computations, PCA and nonparametric ANCOVAs have been performed with R software (v.3.4.2) – packages *corrplot, FactoMineR* and *sm* respectively.

## Results

One-way ANOVA showed significant group-dependence for SI, CV and H. Mean and median results are displayed in Table 2, as well as *post-hoc* results.

**Table 2.** Results from the one-way ANOVA applied to the computed parameters SI, CV, H, and D. Means±SD are given for all groups, or medians [Q1-Q3] if normality test failed. The *p*−value showing group influence and *post-hoc* comparisons are shown in the last lines; significant results are written in bold font.

| Group | SI (s) | CV | H | D |
| --- | --- | --- | --- | --- |
| AD | 1.112 [1.080-1.159] | 0.015 [0.010-0.023] | 0.739±0.145 | 1.624±0.150 |
| PD | 1.082 [1.026-1.150] | 0.034 [0.019-0.056] | 0.685±0.185 | 1.576±0.138 |
| HUNT | 1.102 [1.045-1.203] | 0.072 [0.048-0.096] | 0.669±0.155 | 1.544±0.155 |
| ALS | 1.268 [1.204-1.543] | 0.061 [0.051-0.066] | 0.842±0.176 | 1.596±0.102 |
| $p$ | **<0.001** | **<0.001** | **<0.001** | 0.513 |
| AD *vs* PD | NS | **<0.001** | 0.661 |  |
| AD *vs* HUNT | NS | **<0.001** | 0.616 |  |
| AD *vs* ALS | **0.015** | **<0.001** | 0.371 |  |

A first graphical exploration of Spearman correlations between parameters in Healthy group is given in Fig. 2. SI, CV and H are significantly correlated with age in healthy subjects; detailed values are given in Table 3. As seen in this last table, no correlation with age is observed in pathological subjects but CV is correlated with the disability scales in PD and HUNT groups. H also shows a correlation with H&Y scale in PD group and D is correlated with TFC in HUNT group. Besides correlations shows in Table 3 we notice a significant negative correlation between CV and D in all groups but ALS: Healthy *ρ* = −0.422 (*p* = 0.001), PD *ρ* = −0.400 (*p* = 0.018), HUNT *ρ* = 0.486 (*p* = 0.019).

**Table 3.**
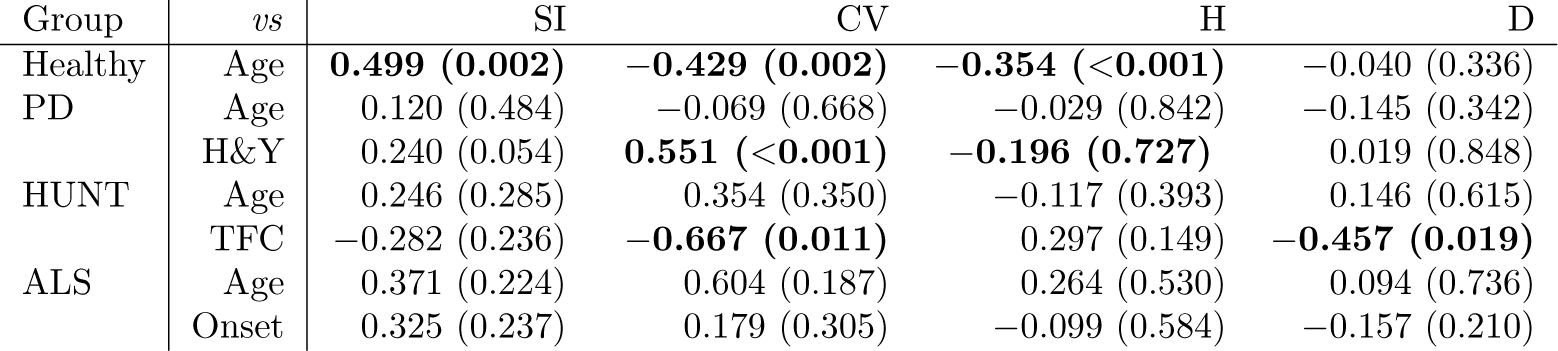
Spearman correlation coefficients between the parameters SI, CV, H, and D and subject’s ages for all groups, and between the different parameters and disability scores for PD (H&Y) and HUNT (TFC) groups, and months since first diagnosis (Onset) for ALS. The *p*−values are given between parenthesis and significant correlations are displayed in bold font.

| Group | vs | SI | CV | H | D |
| --- | --- | --- | --- | --- | --- |
| Healthy | Age | **0.499 (0.002)** | **-0.429 (0.002)** | **-0.354 (<0.001)** | -0.040 (0.336) |
| PD | Age | 0.120 (0.484) | -0.069 (0.668) | -0.029 (0.842) | -0.145 (0.342) |
|  | H&Y | 0.240 (0.054) | **0.551 (<0.001)** | **-0.196 (0.727)** | 0.019 (0.848) |
| HUNT | Age | 0.246 (0.285) | 0.354 (0.350) | -0.117 (0.393) | 0.146 (0.615) |
|  | TFC | -0.282 (0.236) | **-0.667 (0.011)** | 0.297 (0.149) | **-0.457 (0.019)** |
| ALS | Age | 0.371 (0.224) | 0.604 (0.187) | 0.264 (0.530) | 0.094 (0.736) |
|  | Onset | 0.325 (0.237) | 0.179 (0.305) | -0.099 (0.584) | -0.157 (0.210) |

**Fig 2.**
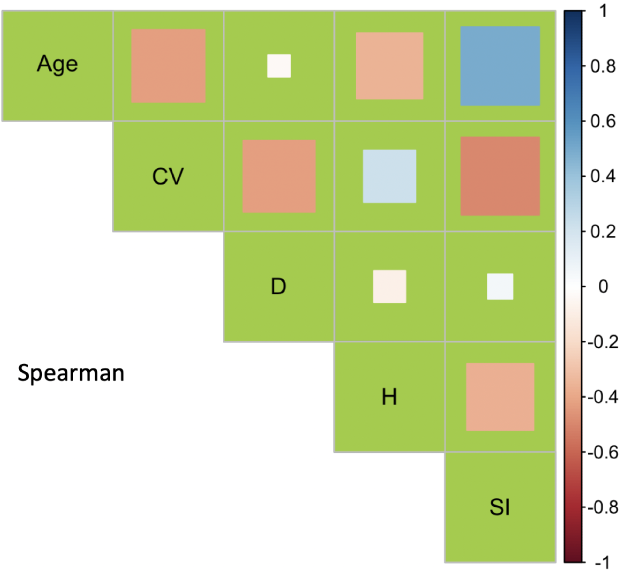
Correlation plot of the Healthy subject’s parameters. Each square has an area proportional to |*ρ*|, the full square being of unit area.

The nonparametric ANCOVA indicates that the behavior of SI *versus* age is different between Healthy and HUNT groups. Healthy and ALS show parallel SI trends with age, shifted by 0.214 seconds upwards in ALS group. In other words, SI of ALS patients are significantly longer than Healthy individuals of the same age. Moreover, the behaviors of CV *versus* age are different between Healthy and HUNT and between Healthy and ALS groups. The trend of H *versus* age are parallel in Healthy and ALS groups, but shifted upwards by 0.176 in the ALS group. Nonparametric ANCOVA was not performed for D since this parameter was not significantly correlated with age even in the Healthy group.

The PCA displayed in Fig. 3 shows that the set of computed parameters (SI, CV,D, H) accounts for 70.1% of the total variability of Healthy group only including the first two dimensions. Moreover the “standard” indices (SI and CV) and the “fractal” ones (D and H) are rather uncorrelated, showing the relevance of including both kind of indices in a characterization of SI time series. Similar conclusions can be drawn from the PCA in the pathological group. We note that CV and SI are negatively (positively) correlated in Healthy (pathological) group.

**Fig 3.**
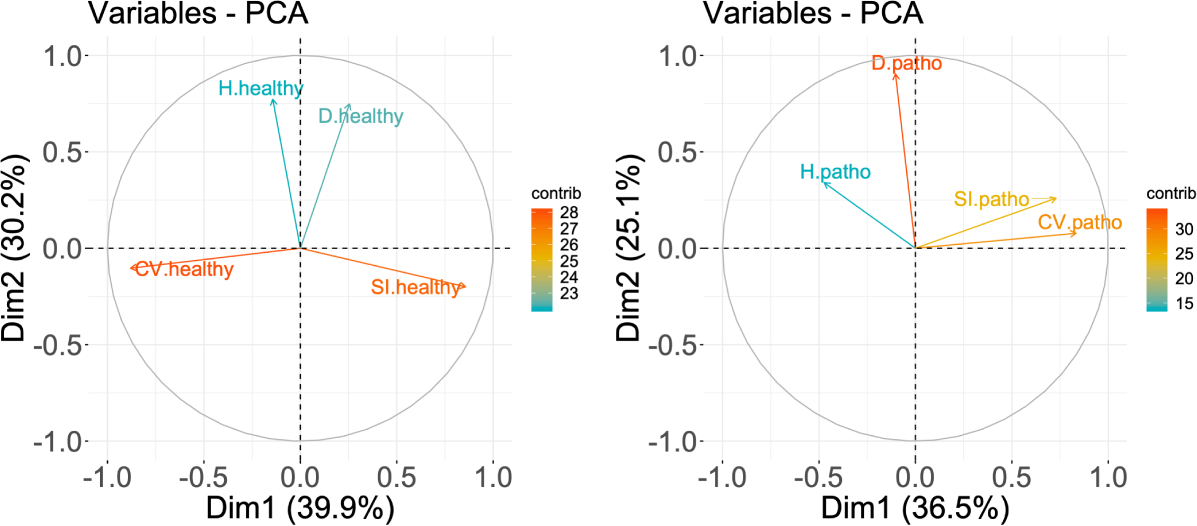
PCA performed on the computed parameters for the Healthy (left panel) and pathological (right panel) groups. The correlation circles for dimensions 1 and 2 are shown. The contribution of each variable to the principal axes (contrib) are coded in colors, with cold colors (turquoise blue) showing low contribution and warm colors (orange) high contribution

Our results are graphically summarized in Fig. 4. Individual results are shown in the pathological groups while a smoothed trend is shown for Healthy group. The most obvious observation is that any pathology tends to increase CV from its healthy value. ALS subjects have a higher SI, as confirmed by the nonparametric ANCOVA.

**Fig 4.**
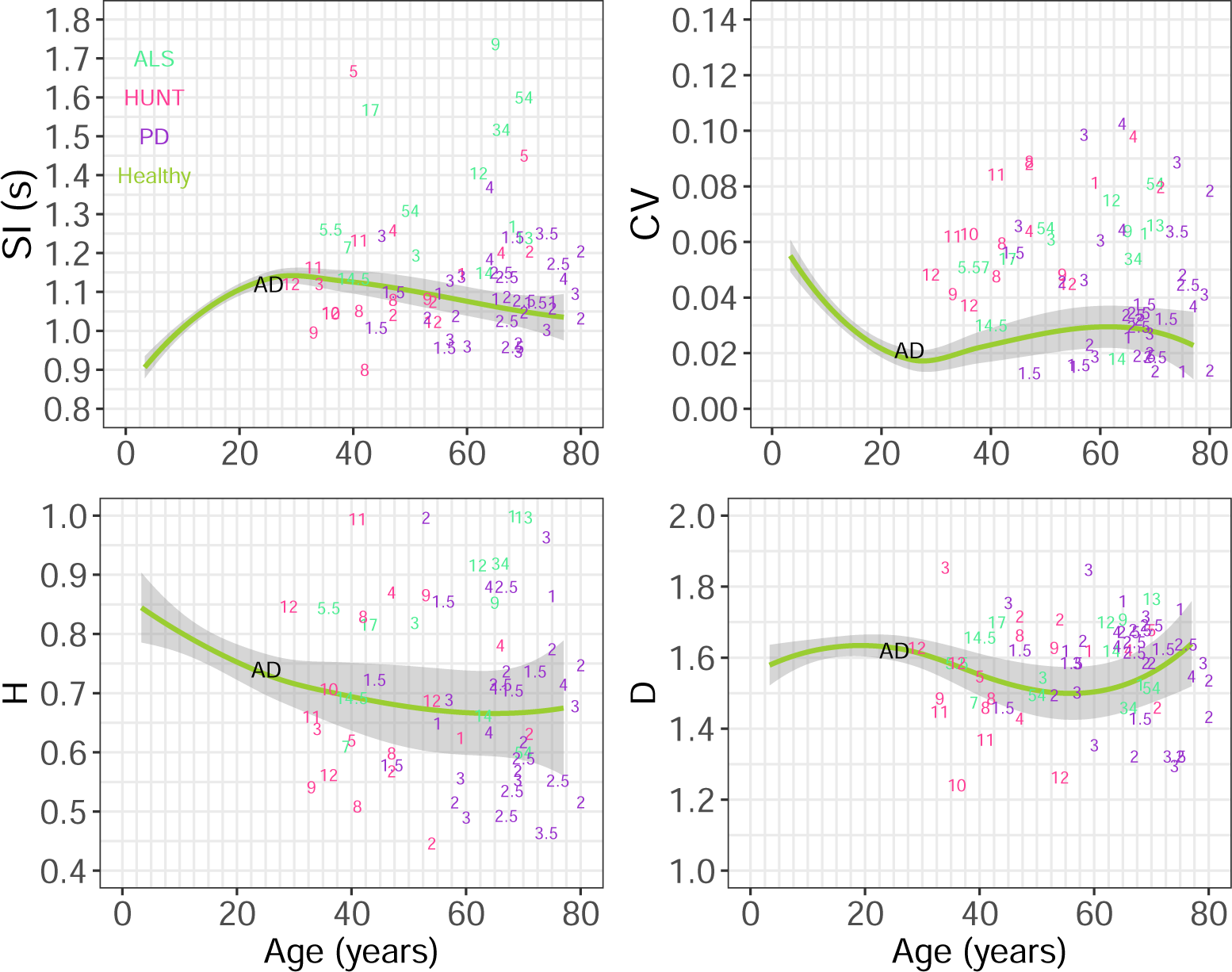
Comparison of pathological subjects (each subject is labeled by the associated disability scores for PD and HUNT, and months since onset of disease for ALS) and Healthy subjects (green solid lines +95% grey confidence intervals) concerning the evolution of the computed parameters SI, CV, H, and D *versus* subject’s age. A label AD is located at the average position of AD group. The Healthy group trends have been obtained by a second-order polynomial smoothing of individual data (*sm* package in R).

## Discussion

The objective of this study was to recompute linear and nonlinear variability indices in a uniform way from raw SI time series collected previously in healthy subjects and three common neurodegenerative diseases. In addition to variability indices computed from SI, the patient’s level of disability was also correlated with these indices. Measures of variability in general are more and more frequently considered in gait analysis in patients with neurodegenerative diseases and their clinical utility makes no doubt. Finding new ways to improve the sensitivity of these methods is therefore highly relevant. Here we consider more advanced yet practically simple indexes such as the Hurst exponent and the Minkowski fractal dimension in an attempt to provide a more accurate clinical description of patients. Our findings show that these indices indeed probe other features of SI time series (see Fig. 3) than usual SI average value and coefficient of variation. Moreover several variability indices were significantly correlated with the disability scores.

In healthy subjects, the average SI is an increasing function of age from birth to approximately 20 years and eventually reaches a plateau. This observation could be partly explained by a mechanical effect of growth: in any pendular model of walk for example, 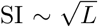 is expected, with *L* the lower limb’s length. Our observations are compatible with well-known results about changes in gait patterns of growing children, see [24]. While growing, children also learn to control their gait, and the magnitude of SI fluctuations decreases: CV reaches a minimal value of 0.015 [0.010 − 0.023] in AD group. This value is compatible with the meta-analysis conducted by Moon et al. [25] reporting an average value of 0.024±0.005 for CV in healthy adults. The healthy adults thus reach an optimal control of their walk; any deviation from this optimal state will result in an increase of CV. Hurst exponent is found to be equal to 0.739±0.145 in AD group. Our value is very close to that of 0.75 found in the one-central pattern generator (CPG) model proposed by Hausdorff et al. [3]. In contrast, the fractal dimension does not significantly vary with age. As detailed in a previous study of our group [13], we interpret this parameter as a complexity index of the SI time series, see [8] for a discussion of this parameter in the case of electrocardiographic time series. According to the maximal complexity model [16], D should be maximal in healthy subjects, as an indicator of optimal adaptability to external environment. Here, we observe no significant variation of D with aging, but we point out the negative correlation obtained between D and CV: healthy adult reaches minimal CV which corresponds to large values of D, in-line with the optimal complexity framework.

Maturation of the nervous system may be seen in electromyographical activity while walking. It has been shown [26, 27] that basic patterns exist from birth and become more and more complex while growing. It can be reasonably assumed that the increase in complexity of walking muscular activation patterns is associated with a better control of walk including SI duration, hence with a lower CV. Simpler activation patterns should also lead to a more stereotyped and predictable walk. Indeed, we observed larger values of H in healthy children compared to healthy adults. An interesting comparison with the biomechanical model of Gates et al. [28] can be made at this stage. The authors use a pendular model of walk, with initial condition updated at each new step, i.e. at each heel strike. Sensory and motor noise, regulated by a simple proportional feedback controller, were incorporated in the model to vary the push-off forces generated by the trailing leg from step to step. Large H values can be obtained for large values of feedback control which makes sense since children’s walk are more consciously controlled. CV and H tend to increase and decrease respectively in healthy old adults with respect to the adult condition. This may simply be related to physiological aging (sarcopenia, joint stiffness, etc.), leading to a less efficient control (higher CV) and a general disorganization of long-range walking variability (smaller H). This goes along the lines of the model discussed in [28], where an increase in motor noise (a random noise added to the feedback controller) systematically leads to the behaviors we observe in old adults.

The existence of long-range autocorrelations in SI variability has always been thought to be rooted in the central nervous system [3], and models involving central pattern generators (CPGs) have been shown to successfully reproduce such autocorrelations [29]. CPGs are sets of neurons located in the spinal cord and capable of regulating an automated rhythmic action. Naturally, several studies have focused on variability of walk in patients suffering from neurodegenerative diseases (for a recent systematic review on this topic see [25] and references therein). Parkinson’s and Huntington’s diseases both lead to an alteration of the basal ganglia, and the degeneration of the central grey nuclei will lead to an alteration in the transmission of data to the CPG via the mesencephalon [30]. Such perturbation necessarily increases gait variability and CV with respect to an healthy adult, as confirmed by the nonparametric ANCOVA (Table 4) in Huntington’s case and by the *post-hoc* results in all pathological cases (Table 2). However, Parkinson’s and Huntington’s diseases have an opposite clinical manifestation: Parkinsonian, lacking of dopamine neurons, suffer from hypokinesia while the patient suffering from Huntington’s disease suffer from hyperkinesia due to a decrease in striatum activating neurons. The H exponent is indeed significantly lower in the HUNT group compared to other pathological groups: hyperkinesia induces random perturbation of the SI, i.e. reduces the regularity or predictability of the lower limb movements. Consequently, from a clinical perspective, H could reflect the decreased dynamic stability of walking kinematics and therefore an increased risk of fall observed in patients with Huntington’s disease [31]. Lyapunov exponents may assess similar features of walking kinematics [32]. A detailed analysis of our findings revealed that, for patients with Parkinson’s disease with low H&Y scores (1 and 1.5), H is higher than in healthy subjects of the same age. It underlines a more predictable or stereotyped walk. In contrast, H is globally lower in patients with Parkinson’s disease with higher H&Y scores than in healthy subjects of comparable age. As for Huntington’s patients, walk becomes more random. From a clinical perspective, since low H&Y scores are mainly observed at early stages of the disease, an increase of H value for SI could help in the early diagnosis of Parkinson’s disease, at a time when it is precisely difficult to diagnose because it is based solely on the symptoms observed. Our findings are in agreement with a previous study [33] that reported a negative correlation between modified H&Y score and Hurst exponents.

**Table 4.** Results of nonparametric ANCOVA comparing the Healthy group *versus* pathological groups. The null hypothesis of equal behaviors of parameters *versus* time is tested in the first line (Equal). The null hypothesis of parallel behaviors of parameters *versus* time is tested in the second line (Parallel). In case this last hypothesis is not rejected, the shift between both groups is given in the third line (Shift). Significant differences are written in bold font.

| Healthy *vs* | SI |  |  | CV |  |  | H |  |  |
| --- | --- | --- | --- | --- | --- | --- | --- | --- | --- |
|  | PD | HUNT | ALS | PD | HUNT | ALS | PD | HUNT | ALS |
| Equal | 0.208 | **<0.001** | **<0.001** | 0.095 | **<0.001** | **<0.001** | 0.936 | 0.663 | **0.004** |
| Parallel | – | **<0.001** | 0.324 | – | 0.132 | **0.063** | – | – | 0.303 |
| Shift |  |  | 0.224 s |  | 0.065 |  |  |  | 0.117 |

In Huntington’s disease, CV is negatively correlated with TFC: the smaller TFC (the more serious the disability), the larger the CV. A correlation between H and TFC was found in [4] but not confirmed by our computations. Note however that we calculate H by using another algorithm. Our results confirm that an alteration of SI variability is linked to a loss of functional ability in patients suffering from neurodegenerative pathologies. Would a treatment aiming at improving a patient’s variability, assessed by CV, H and D, lead to an improvement of his/her functional ability? This question is, to our knowledge, still to be addressed.

ALS is a progressive degeneration of motor neurons in the cerebral cortex and spinal cord. It affects the cortical-spinal pathway and motor neurons of the motor units of skeletal muscle, which will create paralysis of these muscles. A lack of “peripheral” muscle control logically leads to a higher value of CV, and the muscle paralysis is associated to a significantly higher value of SI in ALS group. Quite not significant, patients suffering from ALS also show a higher value of H, exhibiting a more predictable gait pattern.

Some limitations of our study should be mentioned. First, the length of available time series were variable which is known to induce potential biases in the estimation of indexes such as Hurst exponents. To limit the risk of artefacts, we have decided to truncate time series to 256 points (or close to), which may be a minimal length to compute accurate variability indices [20]. Even if the duration of a walking session – around 8 minutes for all the time series of our data set – is a natural parameter, a more rigorous analysis scheme will be reached by standardizing the number of steps in future studies. Second, no disability score was available for ALS patients and no correlation could be calculated with the nonlinear variability indices. Third, Hurst exponents were only computed by using the Detrended Fluctuation Analysis for the sake of homogeneity. However, Rescaled Range Analysis has also been used previously [34] and may have been included also. Finally, other nonlinear analysis tools such as Lyapounov exponents [35] could have been computed in order to consider their added value. However, to our knowledge, they would not provide complementary information but would only confirm how intrinsic properties calculated by other variables such as H vary.

In summary, we have confirmed that healthy adults reach an optimal variability: CV is minimal, D is maximal, and H reaches an optimal value, typical of chaotic systems [16]. Any deviation from this optimal state, either through aging or pathology, will cause CV to increase. Hence, CV is a linear relevant index but we recommend to complement it with nonlinear indices. D is expected to be lower in pathological cases and has been shown to be negatively correlated to CV. A patient showing both larger CV and D values than a healthy adult may be considered as pathological too. Moreover, H is sensitive to H&Y scale in Parkinson’s disease, and significantly lowered in patients suffering from Huntington’s disease. The assessment of non-linear variability indexes must be progressively introduced in clinical practice in order to aid clinical assessment, track disease progression, optimize pharmacological treatment and rehabilitation follow-up in neurodegenerative patients.

## Data Availability

All data referred in the manuscript are available.

## Notes

### Competing Interest Statement

The authors have declared no competing interest.

### Funding Statement

No external funding was received to support this work.

